# Healthcare professionals’ knowledge and practices towards hospital infections in surgical clinics

**DOI:** 10.1101/2021.11.05.21265982

**Authors:** Petros Galanis, Katerina Kokkoliou, Irene Vraka, Olympia Konstantakopoulou, Olga Siskou, Angeliki Bilali, Daphne Kaitelidou

**Author notes:** **Corresponding author:** Petros Galanis, PhD, Faculty of Nursing, Center for Health Services Management and Evaluation, National and Kapodistrian University of Athens, 123 Papadiamantopoulou street, GR-11527, Athens, Greece.

## Abstract

**Background:** Hospital-acquired infections are a major public health problem, as they increase hospitalization, cost, morbidity, mortality and antibiotic resistance.

**Aim:** To assess the level of knowledge and practices of health professionals about hospital infections in surgical clinics and investigate possible determinants that affects their compliance with international protocols for prevention and control of hospital infections.

**Methods:** A cross-sectional study with a convenience sample was conducted. Study population included 106 health professionals from medical and nursing staff in surgical clinics of a general hospital in Attica. Data collection was conducted during October and December 2019. We used the Healthcare-Associated Infections questionnaire to measure knowledge and practices of health professionals about hospital infections.

**Results:** The mean overall knowledge score for hospital infections was 59.4, indicating a moderate level of knowledge. The highest level of knowledge was about the safety of healthcare professionals, while the lowest level was about the source of hospital infections. The results of the multivariate linear regression showed that participants who were aware of the infection control program at their hospital and knew they had to wash their hands following the six steps of the hand hygiene protocol had a higher level of knowledge score regarding hospital infections. In contrast, participants who felt that their hospital was following a good infection control and prevention strategy had a lower level of knowledge. In addition, healthcare professionals who washed their hands before and after examining patients, after using medical equipment for patient care, throughout and after the end of the shift, and after removing disposable gloves had a higher level of knowledge for hospital infections.

**Conclusions:** The findings of the present study are consistent with the international literature on the existence of a moderate level of knowledge regarding international prevention guidelines. Compliance of healthcare professionals is essential to achieve universal, quality and safe healthcare and a safe working environment.

## Introduction

Hospital-acquired infections are a major public health problem, as they increase hospital time and costs, morbidity, mortality, antibiotic resistance and the likelihood of healthcare workers being exposed to infections (Pittet et al., 2008). It is crucial to understand the importance of the existence of hospital-acquired infections, as both the health of staff and patients are at higher risk. Management cost of hospital-acquired infections are not only confined within health structures, but also directly affect society by reducing productivity and family income (Allegranzi et al., 2011; Dimick et al., 2004).

Worldwide, systematic efforts are being made to reduce hospital-acquired infections, but their control is a complex process, with the main obstacle being the non-compliance of health professionals with international guidelines (Haque et al., 2018). Surgical clinics are an additional challenge, since they are closed departments where skilled actions are performed. For this reason, it requires staffing by well-trained personnel with a strongly cultivated climate of adherence to protocols to safeguard patient safety and ensure the provision of optimal health care services (Luo et al., 2010; Nofal et al., 2017). These protocols are about the use of protective equipment, sharps injury avoidance techniques, systematic hand hygiene and the prescribed management of hospital waste during the performance of any nursing procedure (Kriari et al., 2018; Reda et al., 2010; Siegel et al., 2007).

The most important factors that reduce health professionals’ compliance with protocols are lack of knowledge, lack of personal protective equipment, poor quality and poor application of personal protective equipment, burnout due to excessive workload, inability to properly manage emergencies that create stress and tension, lack of supervision of the implementation of protocols, and the absence of a safety climate in to the hospitals (Donati et al., 2019; Hosoglu et al., 2011; Kriari et al., 2018; Luo et al., 2010; Nofal et al., 2017; Reda et al., 2010; Sessa et al., 2011; Wasswa et al., 2015).

The aim of this study was to assess the knowledge and practices of healthcare professionals regarding hospital-acquired infections in surgical clinics and to investigate the factors influencing their compliance with guidelines.

## Materials and methods

### Study design

A cross-sectional study was conducted with physicians and nurses working in the surgical clinics of a general hospital in Athens. Convenience sampling was conducted between October and December 2019. The participation rate was 70.7% (=106/150). We used the Healthcare-Associated Infections questionnaire (Zhou et al., 2014) which has been translated and validated in Greek (Kriari et al., 2018) to collect information. The knowledge scores obtained from the questionnaires take values from 0 to 100% with higher values indicating more knowledge. The outcome in our study was the attitudes and compliance of health professionals regarding hospital-acquired infections in surgical clinics.

The participation of healthcare professionals in this study was voluntary, with no restrictions on the time of completion of the questionnaire and after they had been informed in advance, about the design and purpose of the study. The study protocol was approved by the scientific and ethics committee of the hospital in which the study was carried out (reference number: 348, date of approval: 12/09/2019).

### Statistical analysis

Categorical variables are presented as absolute (n) and relative (%) frequencies, while quantitative variables are presented as mean (standard deviation) or median (interquartile range). The Kolmogorov-Smirnov test and normality plots were used to test the normal distribution of quantitative variables. Bivariate analysis included independent samples t-test, Spearman’s correlation coefficient and Pearson’s correlation coefficient. In case where more than two independent variables were found to be statistically significant at the 0.2 level (p<0.2) in the bivariate analysis, multivariate linear regression was applied with scores as the dependent variable. In this case, the backward stepwise linear regression method was applied. Regarding multiple linear regression, the coefficients b (coefficients’ beta), the corresponding 95% confidence intervals and p-values are presented. The two-sided level of statistical significance was set equal to 0.05. Data analysis was performed using the IBM SPSS 21.0 (Statistical Package for Social Sciences) statistical package for social sciences.

## Results

The study population included 106 health professionals. In particular, 51.9% (n=55) were males and 48.1% (n=51) were females. The mean age was 36.7 years (standard deviation=9.2), and the median number of years of experience was 9 (interquartile range=9.5). 67% (n=71) were medical staff, while 33% (n=35) were nursing staff. The knowledge scores of health professionals regarding infections are presented in Table 1. The mean overall knowledge score was 59.4% indicating a moderate level of knowledge. Higher level of knowledge was about healthcare professionals’ safety and pathogens associated with infections. Moderate level of knowledge was about personal protection, isolation conditions and basic concepts. The level of knowledge about the source of hospital-acquired infections was the lowest.

**Table 1.** Knowledge scores of healthcare professionals regarding hospital-acquired infections.

| **Scale** | **Mean** | **Standard deviation** | **Median** | **Minimum value** | **Maximum value** |
| --- | --- | --- | --- | --- | --- |
| Basic concepts | 60.5 | 13.0 | 58.3 | 16.7 | 83.3 |
| Pathogens associated with infections | 72.6 | 8.1 | 72 | 48 | 92 |
| Source of hospital-acquired infections | 42 | 14.7 | 39.3 | 10.7 | 85.7 |
| Personal protection | 69.3 | 14.6 | 70 | 40 | 100 |
| Healthcare professionals' safety | 83.5 | 29 | 100 | 0 | 100 |
| Isolation conditions | 61.0 | 12.3 | 57.1 | 28.6 | 92.9 |
| Total | 59.4 | 6.7 | 59.8 | 45.7 | 78.3 |

Values are expressed in percentages.

Healthcare professionals’ responses on their practices for the prevention and control of hospital-acquired infections are presented in Table 2. The majority of healthcare professionals reported that they wash their hands with water and liquid soap (92.5%) and wear disposable gloves during clinical examination (86.8%). The situations in which they wash their hands most often are before each meal (73.6%), before and after patient examination (68.9%), between two different procedures on different patients (67.9%) and before performing invasive procedures at the patient’s bedside (62.3%). About of healthcare professionals reported that they cleaned their stethoscope after each patient examination (53.8%), while only 24.5% washed their gown daily. Most healthcare professionals reported that they use a simple surgical mask to transport patients with seasonal influenza (80.2%) and pulmonary tuberculosis (70.5%). 59.4% of healthcare professionals reported that infectious waste from patients is disposed of in a yellow waste cell and 34% in a red waste cell. 70% of physicians reported that they prescribe according to guidelines and protocols. Bivariate relationships between the independent variables and the knowledge score for the source of hospital-acquired infections are presented in Table 3. According to the results of multivariable linear regression, healthcare professionals who knew about the hospital infection control program had more knowledge about the source of hospital-acquired infections (coefficient beta=10, 95% confidence interval=4.6-15.4, p<0.001) and healthcare professionals who thought that their hospital did not follow a good infection control and prevention strategy had more knowledge about the source of hospital-acquired infections (coefficient beta=7, 95% confidence interval=1.2-12.4, p=0.001).

**Table 2.** Healthcare professionals’ responses on their practices for the prevention and control of hospital-acquired infections.

| Question | N (%) |
| --- | --- |
| **How do you usually wash your hands at work?** |  |
| With water | 1 (0.9) |
| With water and solid soap | 5 (4.7) |
| With water and liquid soap | 98 (92.5) |
| With alcohol-soaked tissues | 12 (11.3) |
| **When do you wash your hands?** |  |
| Before each meal | 78 (73.6) |
| Before performing invasive procedures at the patient's bedside | 66 (62.3) |
| Before and after examination of patients | 73 (68.9) |
| Before and after contact with exposed patient skin; with bare hands; unless contact is made with gloves | 50 (47.2) |
| Between two different manipulations on different patients | 72 (67.9) |
| Between two different manipulations on the same patient | 27 (25.5) |
| After using medical equipment to care for patients | 49 (46.2) |
| After using the computer and the desk in the nursing department | 20 (18.9) |
| Throughout and after the end of the shift | 54 (50.9) |
| After removal of disposable gloves | 60 (56.6) |
| **When do you wear disposable (non-sterile) gloves?** |  |
| When using the computer and medical equipment for patient care | 8 (7.5) |
| When prescribing medicines | 2 (1.9) |
| During clinical examination | 92 (86.8) |
| When visiting hospital wards | 46 (43.4) |
| None of the above | 5 (4.7) |
| **How often do you clean your stethoscope with an alcohol solution (e.g. 70% alcohol)?** |  |
| After examining each patient | 57 (53.8) |
| Daily | 15 (14.2) |
| 1 time per week | 5 (4.7) |
| 1 time per month | 11 (10.4) |
| Never | 18 (17.0) |
| **How often do you wash your medical gown or uniform?** |  |
| Daily | 26 (24.5) |
| 3 times per week | 29 (27.4) |
| 2 times per week | 21 (19.8) |
| 1 time per week | 22 (20.8) |
| 1 time per month | 6 (5.7) |
| Never | 2 (1.9) |
| **Have you ever been soiled with blood/vommit or other biological fluids of a patient?** |  |
| Yes | 104 (98.1) |
| No | 2 (1.9) |
| **Have you ever been pierced by a used needle?** |  |
| Yes | 79 (74.5) |
| No | 27 (25.5) |
| **Which patients would you apply a simple surgical mask to during transport to and from the ward for further investigation?** |  |
| In patients with seasonal influenza | 85 (80.2) |
| In patients coughing with suspected pulmonary tuberculosis | 74 (70.5) |
| Patients undergoing radiotherapy for the treatment of colorectal cancer | 38 (35.8) |
| All patients regardless of their condition | 19 (17.9) |
| **Where is infectious waste from patients disposed of?** |  |
| In yellow waste cell | 63 (59.4) |
| In a black garbage bag | 1 (0.9) |
| In a red waste bin | 36 (34.0) |
| I don't know | 6 (5.7) |
| **Have you treated patients with multi-drug resistant hospital pathogens? (only for physicians)** |  |
| Yes | 60 (84.5) |
| No | 11 (15.5) |
| **Your prescribing policy is based on (only for physicians):** |  |
| In your experience | 4 (5.7) |
| The practice followed by your superiors | 13 (18.6) |
| In guidelines and protocols (hospital/national) | 49 (70.0) |
| The information on the package leaflet of the medicines | 4 (5.7) |
| **What is your selection criterion when prescribing antibiotics; (only for physicians)** |  |
| Antimicrobial spectrum | 69 (97.2) |
| Economic cost | 35 (50.0) |
| Pharmaceutical company | 34 (48.6) |
| Generation of antibiotic | 40 (56.3) |

**Table 3.**
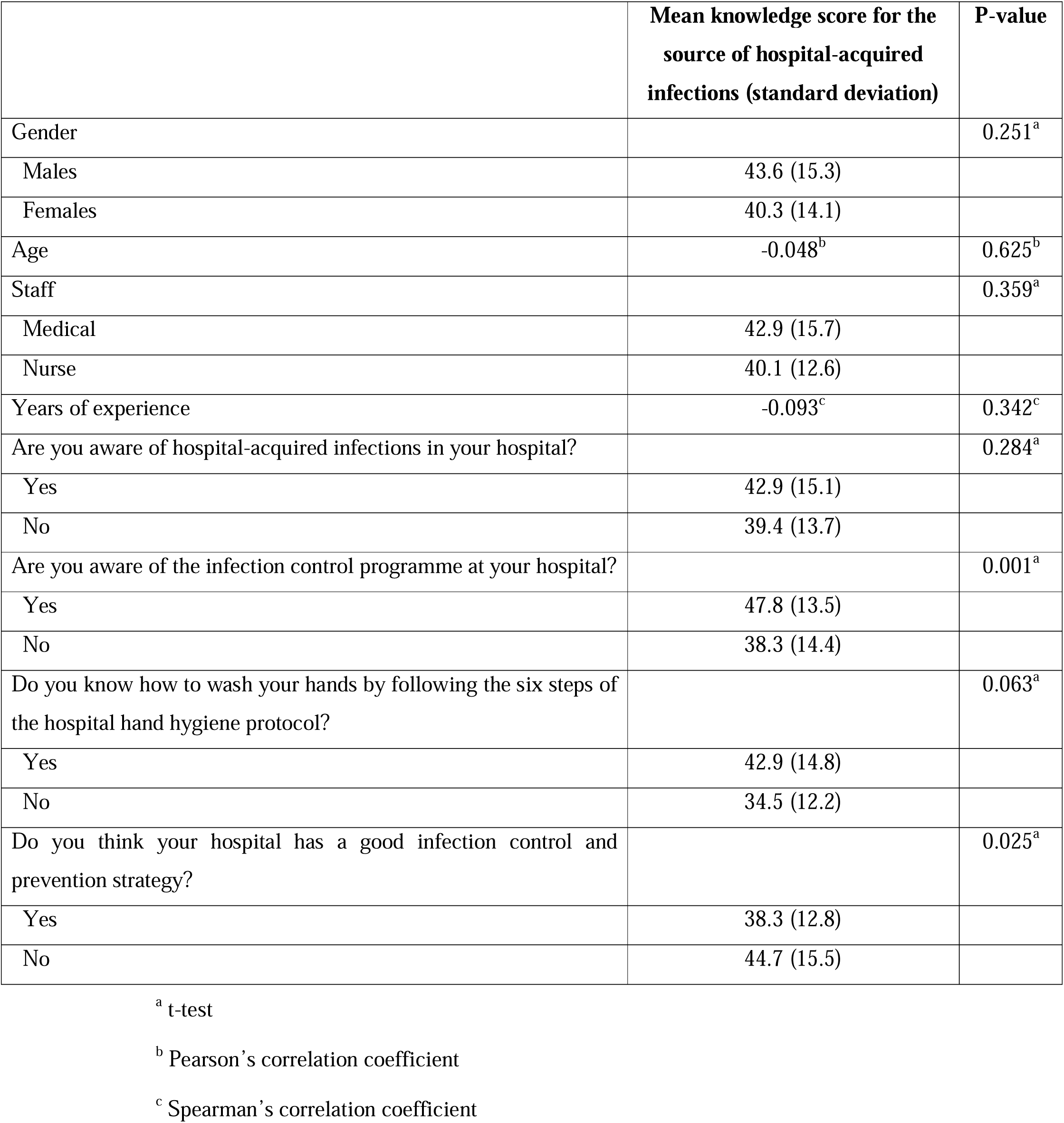
The bivariate relationships between the independent variables and the knowledge score for the source of hospital-acquired infections.

|  | **Mean knowledge score for the source of hospital-acquired infections (standard deviation)** | **P-value** |
| --- | --- | --- |
| Gender |  | 0.251<sup>a</sup> |
| Males | 43.6 (15.3) |  |
| Females | 40.3 (14.1) |  |
| Age | -0.048<sup>b</sup> | 0.625<sup>b</sup> |
| Staff |  | 0.359<sup>a</sup> |
| Medical | 42.9 (15.7) |  |
| Nurse | 40.1 (12.6) |  |
| Years of experience | -0.093<sup>c</sup> | 0.342<sup>c</sup> |
| Are you aware of hospital-acquired infections in your hospital? |  | 0.284<sup>a</sup> |
| Yes | 42.9 (15.1) |  |
| No | 39.4 (13.7) |  |
| Are you aware of the infection control programme at your hospital? |  | 0.001<sup>a</sup> |
| Yes | 47.8 (13.5) |  |
| No | 38.3 (14.4) |  |
| Do you know how to wash your hands by following the six steps of the hospital hand hygiene protocol? |  | 0.063<sup>a</sup> |
| Yes | 42.9 (14.8) |  |
| No | 34.5 (12.2) |  |
| Do you think your hospital has a good infection control and prevention strategy? |  | 0.025<sup>a</sup> |
| Yes | 38.3 (12.8) |  |
| No | 44.7 (15.5) |  |

<sup>a</sup> t-test
<sup>b</sup> Pearson's correlation coefficient
<sup>c</sup> Spearman's correlation coefficient

Bivariate relationships between the independent variables and knowledge score on hospital-acquired infections are presented in Table 4. According to the results of multivariable linear regression, healthcare professionals who knew about the infection control program in their hospital had more knowledge about hospital-acquired infections (coefficient beta=3.8, 95% confidence interval=1.3-6.4, p=0.003), healthcare professionals who knew to wash their hands properly by following the six steps of the hand hygiene protocol in their hospital had more knowledge about hospital-acquired infections (coefficient beta=4.8, 95% confidence interval=1-8.7, p=0.01) and healthcare professionals who felt that their hospital did not follow a good infection control and prevention strategy had more knowledge about the source of hospital-acquired infections (coefficient beta=2.9, 95% confidence interval=0.5-5.4, p=0.02).

**Table 4.**
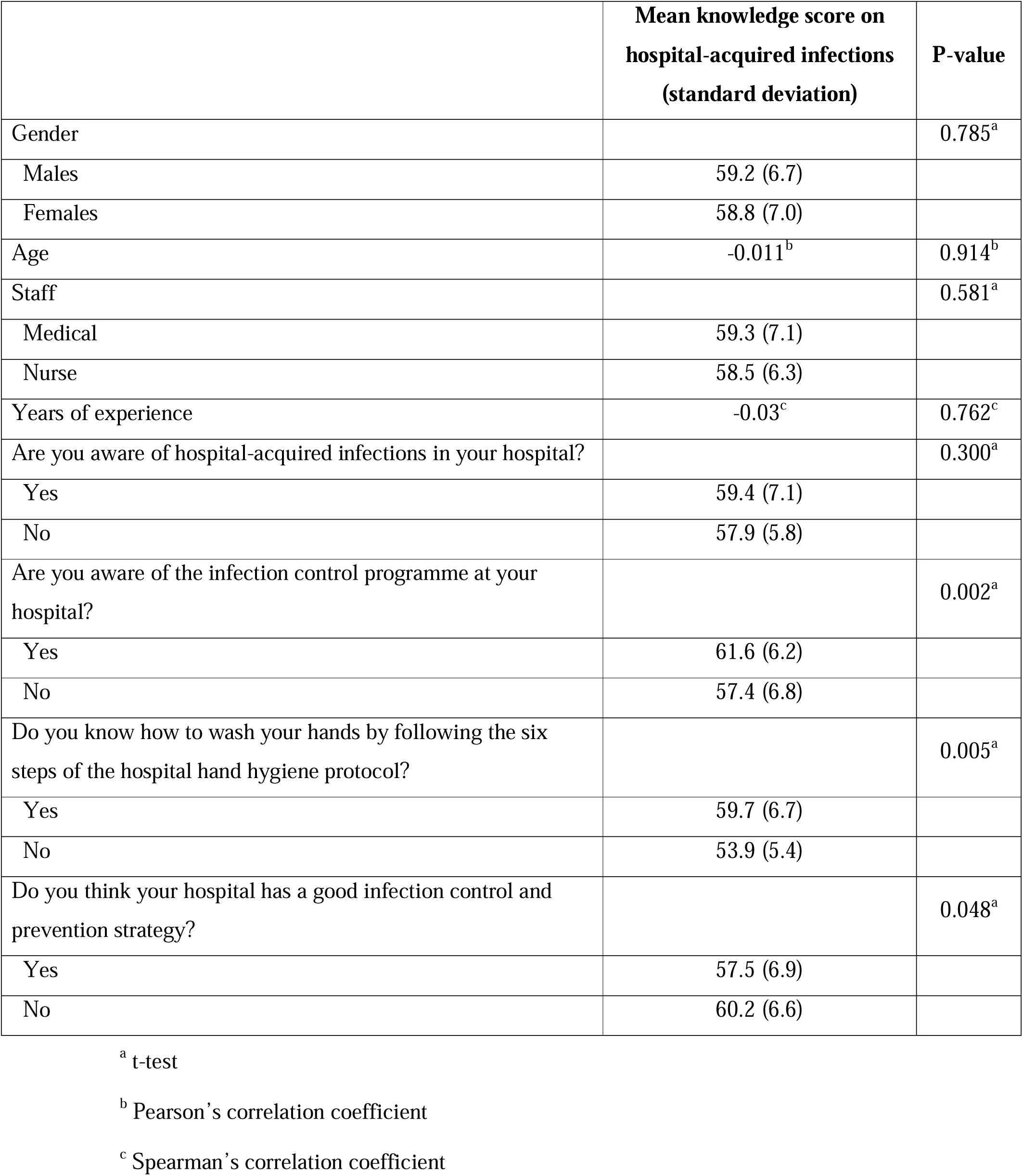
The bivariate relationships between the independent variables and knowledge score on hospital-acquired infections.

|  | **Mean knowledge score on**<br>**hospital-acquired infections**<br>**(standard deviation)** | **P-value** |
| --- | --- | --- |
| Gender |  | 0.785<sup>a</sup> |
| Males | 59.2 (6.7) |  |
| Females | 58.8 (7.0) |  |
| Age | -0.011<sup>b</sup> | 0.914<sup>b</sup> |
| Staff |  | 0.581<sup>a</sup> |
| Medical | 59.3 (7.1) |  |
| Nurse | 58.5 (6.3) |  |
| Years of experience | -0.03<sup>c</sup> | 0.762<sup>c</sup> |
| Are you aware of hospital-acquired infections in your hospital? |  | 0.300<sup>a</sup> |
| Yes | 59.4 (7.1) |  |
| No | 57.9 (5.8) |  |
| Are you aware of the infection control programme at your hospital? |  | 0.002<sup>a</sup> |
| Yes | 61.6 (6.2) |  |
| No | 57.4 (6.8) |  |
| Do you know how to wash your hands by following the six steps of the hospital hand hygiene protocol? |  | 0.005<sup>a</sup> |
| Yes | 59.7 (6.7) |  |
| No | 53.9 (5.4) |  |
| Do you think your hospital has a good infection control and prevention strategy? |  | 0.048<sup>a</sup> |
| Yes | 57.5 (6.9) |  |
| No | 60.2 (6.6) |  |

<sup>a</sup> t-test
<sup>b</sup> Pearson's correlation coefficient
<sup>c</sup> Spearman's correlation coefficient

## Discussion

In this study, the knowledge and practices of healthcare professionals regarding hospital-acquired infections in surgical clinics were investigated. According to the results of multivariate analysis, no relationship was found between demographic characteristics and healthcare professionals’ knowledge and practices. This finding is confirmed by the literature (Chan et al., 2002; Ganczak & Szych, 2007; Kriari et al., 2018; Parmeggiani et al., 2010).

The mean overall knowledge score for hospital-acquired infections in this study was 59.4%, which indicates that both physicians and nurses had a moderate level of knowledge which is in line with similar studies (Chan et al., 2002; Nofal et al., 2017; Sessa et al., 2011; Stein et al., 2003; Wasswa et al., 2015). However, there was a promising conclusion as almost half of the participants (43.4%) reported that the most important method for preventing hospital-acquired infections is the education of healthcare professionals which indicates that they recognize the importance of this knowledge and the need to participate in training programs in order to have an upgrade of knowledge. These results are confirmed by studies in which healthcare professionals recognize the importance of participating in seminars that will help to enhance their knowledge and attitudes to prevent hospital-acquired infections (Donati et al., 2019; Luo et al., 2010; Nofal et al., 2017; Sessa et al., 2011). It has also been found that staff who participate in training programmes show higher levels of compliance with guidelines in clinical practice (Chan et al., 2002; Donati et al., 2019; Luo et al., 2010; Parmeggiani et al., 2010; Wasswa et al., 2015). Furthermore, the absence of relevant educational programmes creates a lack of provision of appropriate information and education and promotion of the importance of implementing the protocols, leading staff to adopt incorrect behaviours, which pose risks to both themselves and patients.

In addition, we found that healthcare professionals who were not aware of the hospital infection control programme had less knowledge. According to the literature there is a wide variation in the level of knowledge of health professionals. Two similar studies found that staff with a higher level of education had more knowledge regarding infections (Nofal et al., 2017; Sessa et al., 2011). In some studies (Kriari et al., 2018) physicians had more knowledge compared to nurses while other studies (Gershon et al., 1995; Parmeggiani et al., 2010) found the opposite conclusion. In addition, one study (Iliyasu et al., 2016) found that knowledge regarding infection control was greater among surgical department workers. This finding may be due to the fact that all techniques followed within the operating room are strictly based on aseptic technique and also on close observation of the patient during recovery to ensure that the surgical wound is not infected.

We found that participants who knew how to wash their hands properly according to the guidelines also had more knowledge for hospital-acquired infections. This finding is confirmed by similar studies that evaluated compliance with international infection prevention guidelines (Gershon et al., 1995; Kriari et al., 2018; Luo et al., 2010; Nofal et al., 2017). Furthermore, 92.5% of the participants reported using water with liquid soap as a method of hand hygiene during working hours, which is the most appropriate method. Considering the above results and the fact that the practice regarding hand hygiene is the main means of controlling and preventing the transmission of hospital-acquired infections, it is noted that there is a fairly good compliance rate among the participants in our study.

It should be noted that knowledge level is not a necessary reason for compliance with the guidelines and therefore there is no significant relation between knowledge level and compliance with international protocols. For example, several studies found that participants with a high level of knowledge and a positive attitude regarding the guidelines for the prevention and control of hospital-acquired infections had a low rate of compliance (Kermode et al., 2005; Ogoina et al., 2015; Parmeggiani et al., 2010; Sessa et al., 2011; Tenna et al., 2013; Wasswa et al., 2015). Moreover, in other studies, although health professionals have at least basic information and knowledge about the guidelines for the control and prevention of hospital-acquired infections and are aware that adherence to them would significantly reduce the pathogens that cause them, their compliance is selective and is estimated to be below desirable levels (Chan et al., 2002; Gershon et al., 1995; Hosoglu et al., 2011; Iliyasu et al., 2016; Luo et al., 2010; Nofal et al., 2017; Ogoina et al., 2015; Reda et al., 2010; Sessa et al., 2011). We found that most healthcare professionals use disposable gloves during clinical examination. Regarding gloves, while there is a good level of knowledge about their proper management, there is a wide variation in the choice of glove use (Kennedy et al., 2004). Several studies report that compliance with glove use is predominantly high only when high-risk cases, such as HIV patients, are to be managed, and predominantly staff does not always use them (Ganczak & Szych, 2007; Gershon et al., 1995; Parmeggiani et al., 2010). According to the guidelines for the prevention and control of hospital-acquired infections, infectious waste such as used syringes, or other used medical equipment should be disposed of in the special red and yellow bags. Various reasons lead to reduced compliance by health care staff, such as the fact that special waste bins are not available in several of the care facilities either because they were not considered necessary in the specific premises or due to lack of appropriate site layout (Wasswa et al., 2015).

Our cross-sectional study has several limitations. Initially the data relates to a given point in time at which the questionnaire was completed. The results of this study were the product of self-completed questionnaires, so information bias is introduced as the responses are likely to be biased. In addition, a convenience sample was used which introduces selection bias. Also, the results are not representative of all health professionals as the study population came from only one hospital in Attica, namely the surgical clinics.

In conclusion, a good level of compliance of the participants regarding the guidelines for the prevention and control of hospital-acquired infections was observed despite a moderate level of knowledge. The relevant health authorities should pay special attention to investigate the factors that influence the attitudes and compliance of health personnel in the prevention of hospital-acquired infections by conducting more studies with a larger number of participants in order to formulate appropriate interventions. In addition, one of the primary objectives of healthcare systems is to provide quality and safe care, in which the reduction of hospital-acquired infections, which are an indicator of the quality of health services provided, plays a key role, as their existence indicates a failure to provide protection against infectious agents.

## Data Availability

All data produced in the present study are available upon reasonable request to the authors

